# Modelling the impact of control measures against the COVID-19 pandemic in Viet Nam

**DOI:** 10.1101/2020.04.24.20078030

**Authors:** Thu-Anh Nguyen, Quoc Nguyen Cuong, Anh Le Thi Kim, Huyen Nguyen Nguyen, Thao Nguyen Thi Huong

**Affiliations:** Woolcock Institute of Medical Research, Hanoi, Vietnam; Sydney School of Medicine, The Faculty of Medicine and Health, The University of Sydney, Australia; Save the Children International, Vientiane, Laos; Hanoi University of Public Health, Hanoi, Vietnam; National Hospital for Tropical Diseases, Vietnam; Strategic Consultancy Company, Hanoi, Vietnam

**Keywords:** SARS-CoV-2, modelling, measures, Vietnam

## Abstract

**Objectives:** Health care system of many countries are facing a surging burden of COVID-19. Although Vietnam has successfully controlled the COVID-19 pandemic to date, there is a sign of initial community transmission. An estimate of possible scenarios to prepare health resources in the future is needed. We used modelling methods to estimate impacts of mitigation measures on the COVID-19 pandemic in Vietnam.

**Methods:** SEIR model built in the COVIDSIM1.1 tool was adopted using available data for estimation. The herd immunization scenario was with no intervention implemented. Other scenarios consisted of isolation and social distancing at different levels (25%, 50%, 75% and 10%, 20%, 30%, respectively). Outcomes include epidemic apex, daily new and cumulative cases, deaths, hospitalized patients and ICU beds needed.

**Results:** By April 8, 2020, there would be 465 infected cases with COVID-19 in Viet Nam, of those 50% were detected. Cumulatively, there would be 1,400 cases and 30 deaths by end of 2020, if 75% of cases was detected and isolated, and 30% of social distancing could be maintained.

The most effective intervention scenario is the detection and isolation of 75% infected cases and reduction of 10% social contacts. This will require an expansion of testing capacity at health facilities and in the community, posing a challenge to identify high-risk groups to prioritized testing.

**Conclusions:** In a localized epidemic setting, the expansion of testing should be the key measure to control the epidemic. Social distancing plays a significant role to prevent further transmission to the community.

## Introduction

The first SARS-COV-2 case was reported from Wuhan, China in December 2019, and has now spread out to 211 countries, territories, and 2 cruise ships with 1,353,361 infections and 79,235 deaths by 8 April 2020. [1] Since 11 March 2020, World Health Organization (WHO) has declared SARS-CoV-2 outbreak a global pandemic.

According to WHO, R_0_ (R naught - the reproduction number), the average number of secondary cases attributable to infection by an index case after that case is introduced into a susceptible population, is estimated at 2.0 to 2.5. Viet Nam is considered successfully controlling the first stage of SARS-CoV-2 outbreak with 16 cases detected in connection with Wuhan. On 22 January 2020, the first COVID-19 case who returned from Wuhan was detected. Other cases afterwards either were related to returning from China or got infections from cases returning from China. By 13 February 2020, all these 16 cases were recovered and discharged from hospitals. However, from 2 March 2020, some passengers on the international flights to Viet Nam were found to be infected with SARS-CoV-2 and transmitted the virus to some of their close contacts. Some clusters were reported from Bach Mai hospital, Truong Sinh company in Hanoi, and Buddha bar in Ho Chi Minh city. By 8 April 2020, Viet Nam has reported 252 confirmed cases, of which 128 cases were recovered, 124 have been treated at health facilities, and no death reported. [2]

Viet Nam has been implementing a number of rigorous interventions to control the COVID-19 pandemic, including school closure; contact tracing, isolation and testing of suspected and probable cases; isolation and treatment of confirmed cases; testing expansion; cancellation of flights from countries of epicenters; and social distancing.

On one hand, these intervention strategies made Viet Nam reduce new cases. On the other hand, these rigorous interventions, if implemented in a long-term, could lead to negative consequences for society and economy in the future, such as, the increase of stress and anxiety, restriction of production, limited business and economic development. These consequences could become more serious for the poor and other vulnerable populations.

In order to achieve comprehensive information in decision making for the next steps of pandemic control, we used modelling methods to estimate impacts of different mitigation measures on the pandemic. We also predicted resources of health system that needed for different pandemic scenarios.

## Methods

The *COVIDSIM1*.*1*, a projection tool developed by ExploSYS GmbH, was used. [3] The tool adopted the SEIR (Susceptible - Exposed - Infectious - Recovered) model that has been widely used for simulating spread of communicable diseases. In this model, the population is categorized into 4 groups: susceptible, exposed, infected groups (symptomatic and asymptomatic), and a group with outcomes (recovery or death). This model was used to assess the effectiveness of the interventions for COVID-19 pandemic in New Zealand and one hypothetical European country. [4] [5] Details of the model and its algorithms are presented in the *Annex 1*.

The *Annex 2* displays input data of the model. Key assumptions include a latency period of 4 days, a prodromal period of 1 day, a fully infective period of 10 days, infections leading to sickness of 82%, and R0 of 3.4 for worst case scenario and 2.5 for other intervention scenarios. Average duration of hospitalization was 15.4 days without critical care and 23.5 days with ICU. We assumed that 30% of sick patients are hospitalized and 10% require intensive care. These assumptions were developed using the data of Viet Nam that are accessible on the website of the Ministry of Health (MOH). Where local data is not available, we have used and adjusted other clinical parameters from other countries.

We assumed that different measures, such as, contact tracing, isolation and testing of suspected and probable cases; isolation and treatment of confirmed cases would continue being implemented in a long-term. Therefore, we modelled two main intervention strategies (i) contact tracing and case isolation, and (ii) social distancing. Details are:

- Scenario 1- Current scenario: isolation of infected and probable cases was implemented at 75% and social distancing to reduce social contacts by 30% as of the second week of the 29^th^ March 2020. [6]
- Scenario 2- Relaxed social distancing based on scenario 1: remain 75% isolation of infected and probable cases and 10% reduction of social contacts.
- Scenario 3- Herd immunization strategy: there was no intervention. In this scenario, we assumed that mass media could have effects on people’s behaviors when the number of cases and deaths increased. This could lead to a reduction of 5% of social contacts (or 95% social contacts would remain).
- Other scenarios: interventions of isolation and social distancing would be implemented at different levels, for instance, isolation of infected and probable cases at 25%, 50%, 75% and social distancing to reduce social contacts 10%, 20%, 30%. Assumptions to develop these intervention strategies considered the feasibility in terms of capacity and resources of health system and people in the community.

The key outcomes of these models consisted of the numbers of new cases per day, cumulative cases, deaths, hospitals and ICU beds required. All these outcomes of different models were compared with each other to identify the best model for current capacity of health system.

## Results

### Application of the SEIR model to develop the current scenario

By using a current scenario that R_0_ is 2.5, 75% of cases is detected and isolated right after disease confirmation, and 30% of social distancing is implemented, we estimated that by April 8, 2020, there would be 465 infected cases with SARS-CoV-2 in Viet Nam, of which 369 cases would be symptomatic (Figure 1). Number of cumulative cases by end of 2020 would be 1,400 with a total of 30 COVID-19 related death. There would be about 3 new cases in the community each day. The Figure 2 showed the reported daily number of new cases from 2 groups: cases imported from other countries and local transmission cases. This figure indicated that Viet Nam has controlled successfully the source of imported infection and the number of local transmission cases has been significantly decreased. The estimated number of new cases, patients and ICU beds would be low and not beyond the ability of healthcare system (Table 1).

**Table 1:** Estimated new case, cumulative infection and death, hospitalization and ICU beds required of proposed scenarios

|  | Scenario 1 | Scenario 2 | Scenario 3 | Scenario 4.a | Scenario 4.b | Scenario 5.a | Scenario 5.b |
| --- | --- | --- | --- | --- | --- | --- | --- |
| **Intervention level** |  |  |  |  |  |  |  |
| % case isolation | 75% | 75% | 0% | 25% | 25% | 50% | 50% |
| % social contact reduction | 30% | 10% | 5% | 10% | 20% | 10% | 20% |
| **Results** |  |  |  |  |  |  |  |
| Epidemic peak | 3/2020 | 3/2020 | 6/2020 | 10/2020 | 11/2020 | 3/2021 | 2022 |
| Number of new cases per day |  |  |  |  |  |  |  |
| - End of 6/2020 | 4 | 12 | 2.6M | 10,500 | 2,600 | 470 | 130 |
| - End of 12/2020 | 3 | 12 | 0 | 10,800 | 470,000 | 140,000 | 3,200 |
| - Epidemic peak | 18 | 20 | 2.9M | 1.3M | 1M | 1M |  |
| Number of cumulative cases by end of 2020 | 1,400 | 3,930 | 91.4M | 76.8M | 61M | 3.7M | 186,000 |
| Number of cumulative deaths by end of 2020 | 30 | 90 | 3.2M | 1.9M | 1.2M | 44,000 | 3,300 |
| Number of required hospitalized cases at the peak | 50 | 50 | 10M | 4.2M | 3.2M | 3.2M | 10,000<sup>†</sup> |
| Number of ICU beds at the peak | 7 | 10 | 1.4M | 0.6M | 0.5M | 0.5M | 1,500<sup>†</sup> |

<sup>†</sup> Data by the end of 2020

**Figure 1:**
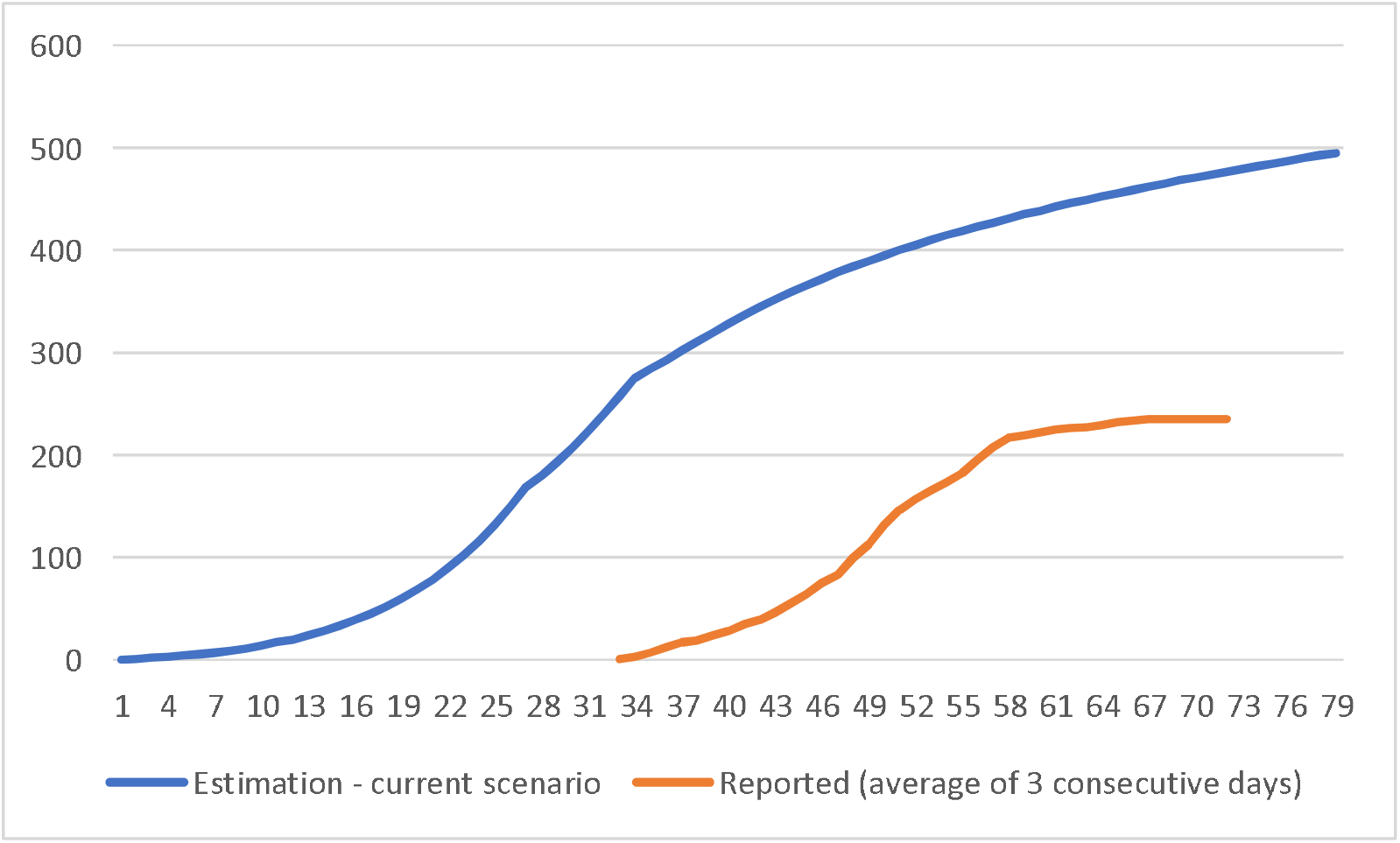
Number of estimated versus reported cases of COVID-19 in Viet Nam

**Figure 2:**
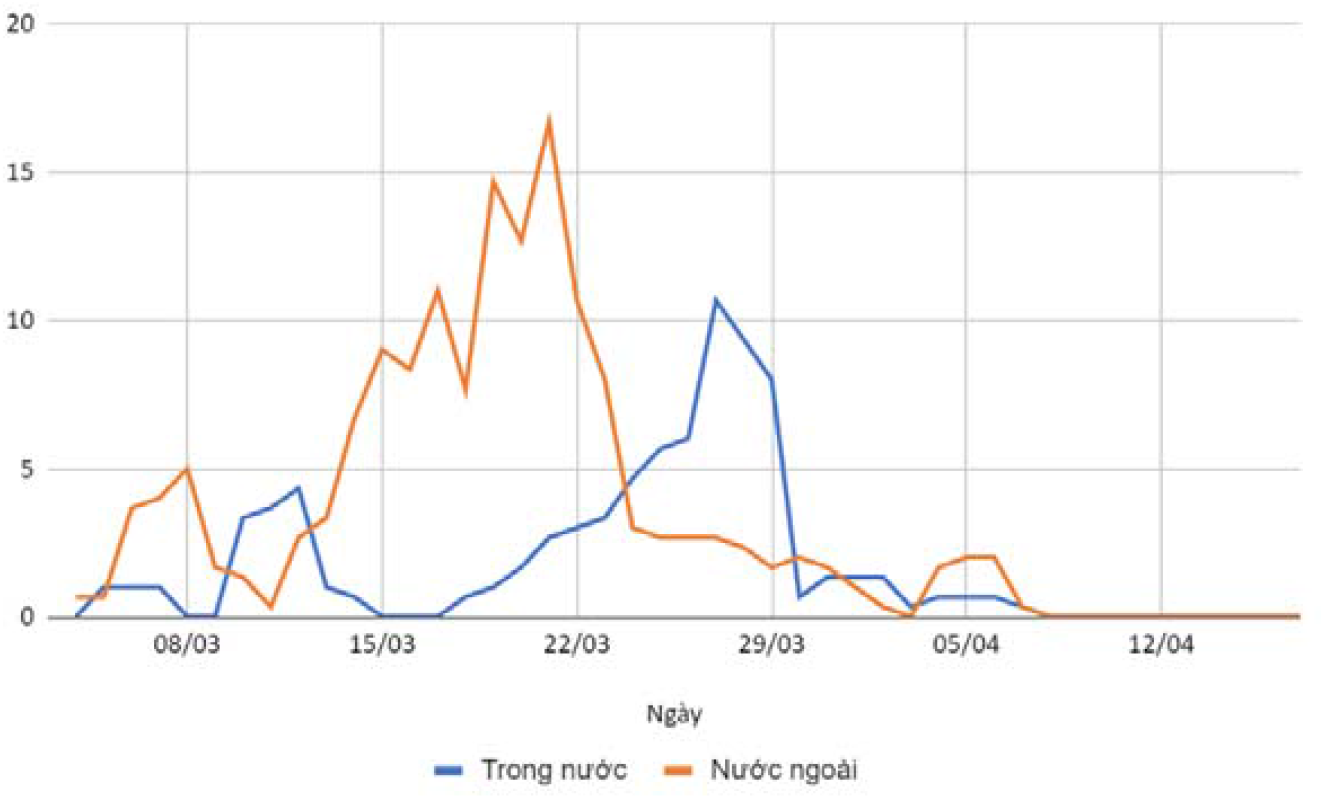
Number of reported and confirmed cases* by day of immigration or day of testing (whichever earlier) ^*\**^*The number of cases per day is calculated as the average of the 3 consecutive days to adjust for delay of reporting time (if any)*.

As presented in the Figure 1, the number of detected cases would be 231, accounts for around 50% of the estimated cases. There are two possible reasons to explain the difference. First, there was a period of no isolation at quarantine centers for immigrants – especially from the US and Europe – to Viet Nam. The regulation of isolation for those from these areas had been released about mid of March while the first case was detected in late January in the US and at the early March 2020 in the Europe. Thus, there could be latent infected cases in the community. Second, there were some cases infected from index cases in the community but they had not been isolated immediately. Moreover, the implementation of contact tracing for confirmed and suspected cases could be incomplete, so their close contacts had been lost of follow up. All these problems could lead to uncontrolled latent cases in the community. In fact, Viet Nam has reported some cases with unknown source of infection.

### Modelling the impact of COVID-19 control measures

Under the herd immunization scenario (scenario 3), we assumed that the Viet Nam Government did not implement any mitigation measures, the number of newly infected cases would peak at 2.9 million per day in June 2020. The number of cumulative cases by the end of 2020 would be 91.4 million, accounting for 95% of total population. The total number of deaths would be 3.2 million, about 2.4% of total population. Figure 3 shows the projection of newly daily cases under scenarios with different level of case isolation and social distancing. Figure 4 indicates the estimated ICU beds required under six scenarios. However, this scenario certainly will not occur since Viet Nam has implemented rigorous and definitive interventions from the outset of the epidemic.

**Figure 3:**
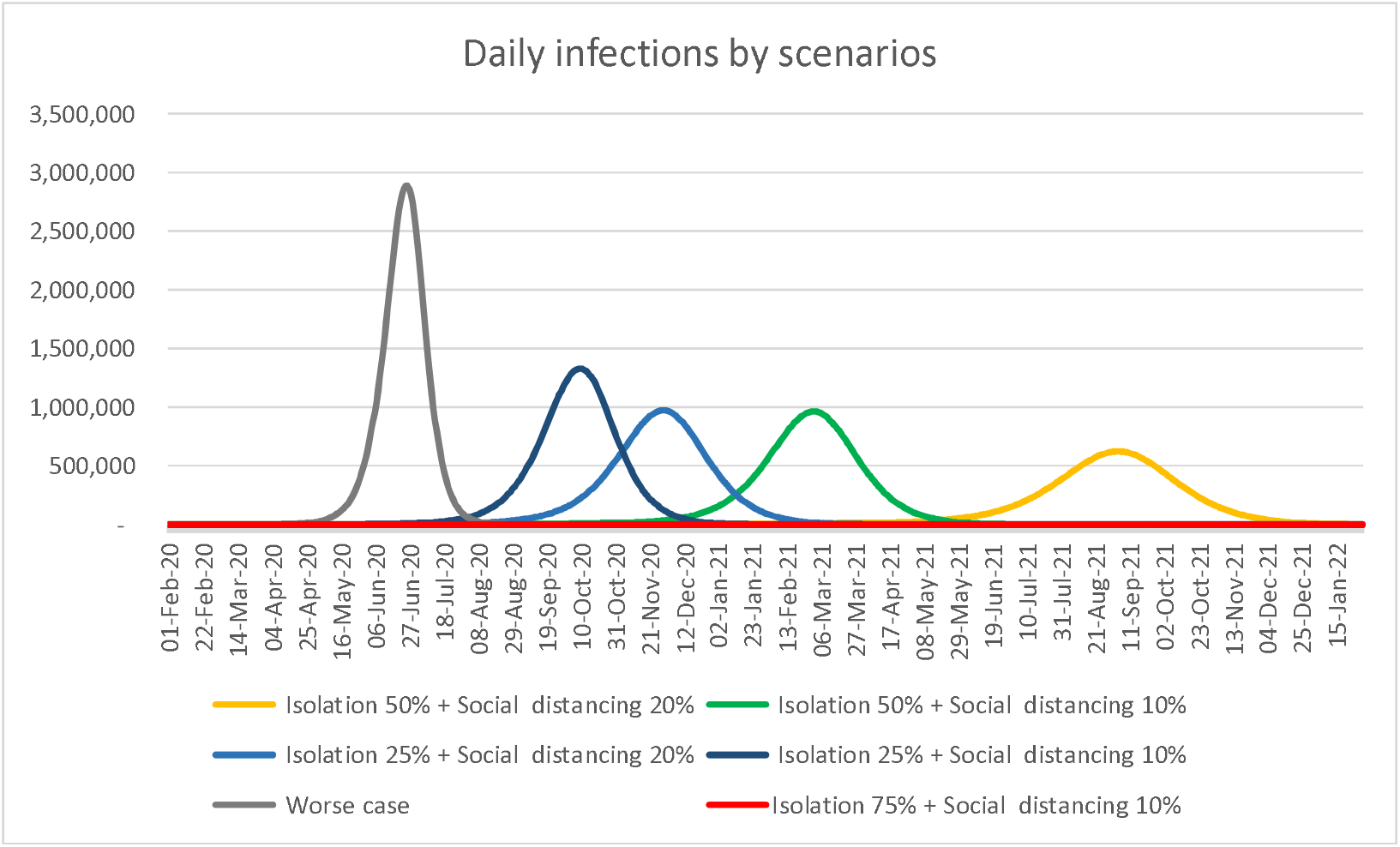
Estimated number of newly daily infected cases by different scenarios

**Figure 4:**
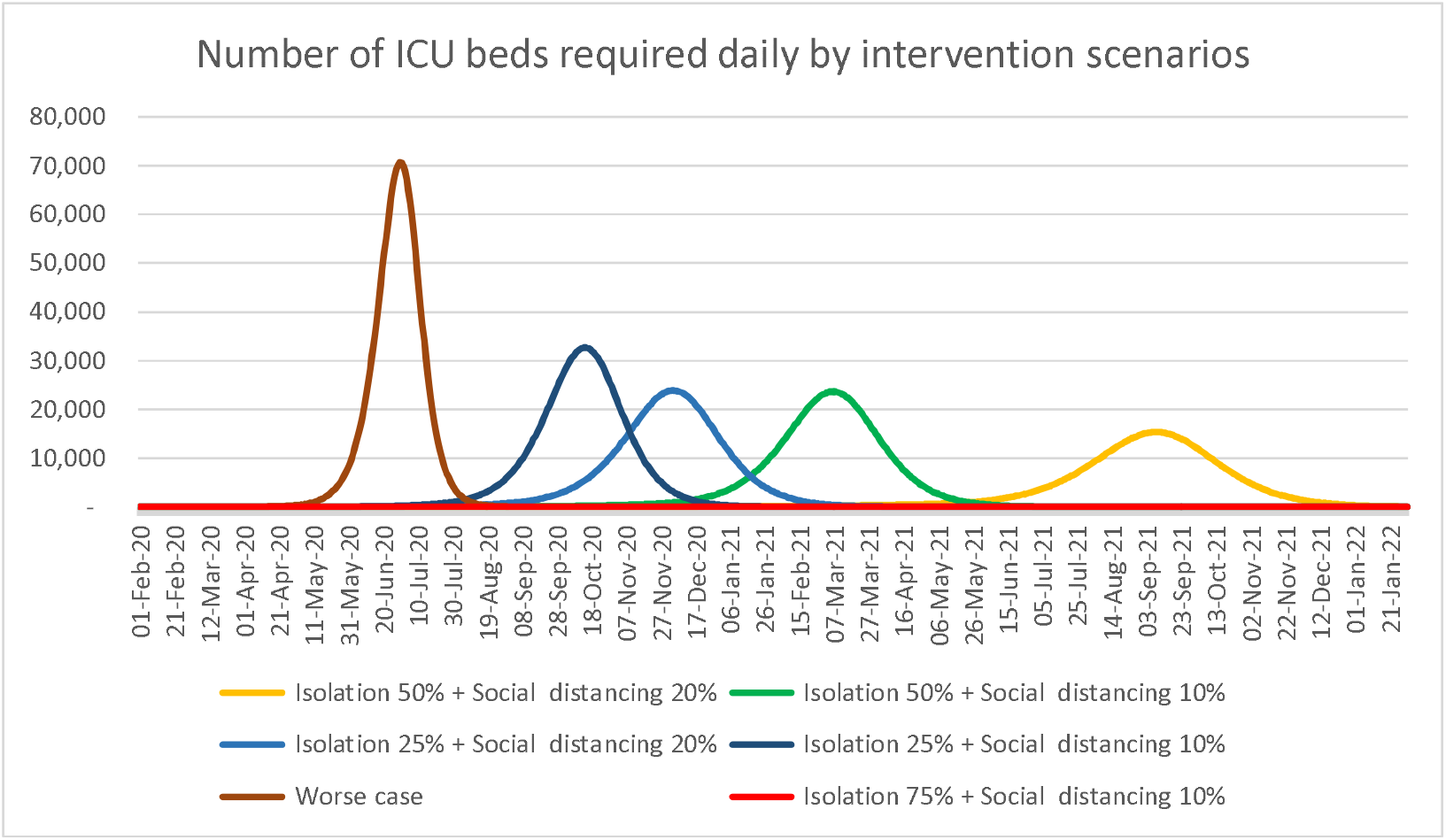
Estimated number of daily new ICU cases by the scenarios

If 25% of infected cases would be isolated, and social contacts would decline by 10% or 20% (scenario 4a and 4b, respectively), the number of new cases per day will be very high, ranging from 1 million - 1.3 million at epidemic peak. The number of ICU beds needed at the peak will be 500,000 to 600,000. In this case, Viet Nam health system (e.g. human resources, ventilators, and infrastructure) will not be able to deal with this demand.

If 50% of infected cases would be isolated, and social contacts would decline by 10% or 20% (scenario 5a and 5b), the pandemic would last and reach its peak by March and September 2022, respectively. Under the scenario 5b, vaccine and effective medication may be available by that time. With this scenario, 1,500 ICU beds and 10,000 patient beds should be prepared by the end of 2020.

If we detected and isolated 75% of confirmed cases and reduced 10% of social contacts, there would be around 12 new cases per day. The cumulative number of deaths by the end of 2020 would be 90. In this scenario, the disease would continue to be endemic in the community with low prevalence.

## Discussion

The results showed that if there was no intervention (scenario 3) or if only 25% of cases detected and quarantined (scenario 4), Viet Nam will experience huge numbers of infections, required IDU beds and deaths in the near future.

In scenarios of isolation of 25% or 50% infected cases, in combination with social distancing of 10% or 20% social contacts, the total number of infected cases in two years (2020-2021) would range from 60%-80% of total population. The raw mortality rate would be 1.5%-1.8%. In other words, these scenarios would save 1.5-1.7 million of lives compared to the worst scenario. If there will be an effective vaccine or medication within this period, the number of infected cases and deaths will be much lower.

The first scenario which is considered to be the current strategy of Viet Nam with rigorous intervention measures are being implemented and an assumption that these measures will last for at least one year would lead to low numbers of infections, required ICU beds and deaths. These intervention measures include detection and isolation of 75% infected cases, and social distancing strategies to reduce social contacts 30%. This scenario, in fact, will require the expansion of testing, not only at health facilities but also in the community to achieve the detection proportion of 75%. Korea, for instance, had done testing expansion, isolation and treatment of SARS-CoV-2 positive cases and the country has got success in reducing the number of new cases and deaths as well. However, the expansion of testing will be a challenge for health system. On average, Viet Nam has detected 1 case among 440 tests in the context of the existence of imported cases that have been under control. It will be a significant challenge to implement this testing strategy because of the required high cost, sufficient testing capacity, infrastructure, and procurement capacity, and ability to provide biological products and consumable supplies to large scale testing. However, if we cannot detect all cases, the probability of detection, and isolation as well, will not reach 75% cases, and the outbreak will then return in the community. Therefore, we might need to combine different approach such as assessing to identify high-risk groups to test them through strengthening the infectious disease surveillance system including influenza like illness surveillance at pharmacy stores, health clinics and hospitals. People presenting at health clinics and hospital with respiratory symptoms should receive a test for SARS-CoV-2.

In addition to expansion of testing, contact tracing up to three generation, and case isolation should be implemented rigorously. People who have close contact with a confirmed case should be isolated for a minimum of 14 days regardless of their test result. The next generation of close contact case should be prepared for contact management procedure if any person in the chain becomes positive. [7]

In addition, the scenario with strict and prolong social distancing to reduce a large number of social contacts would have negative effects on the economic development and social life. The second scenario though will bring the same challenge of an effective testing strategy to detect and isolate 75% of infected cases, it would have a more relaxed social distancing strategy which would reduce only 10% of social contacts. The number of cases and number of required ICU beds in this scenario would be still manageable. This scenario supports strategies to categorize the risk areas and businesses, gradually re-open the economy and social activities in Viet Nam in coming months.

Our study has a number of limitations. So far, we still have limited knowledge of epidemiology and pathology of the SARS-CoV-2. Data of our models were based on multiple assumptions from data of countries where outbreaks have occurred. These models did not adjust for levels of virus transmission that could be affected by weather factors. In fact, we do not have evidences of changing levels of transmission by outdoor temperature. These models did not consider of unusual epicenters that their frequency and context of social contacts could be different from common contacts, such as the recent outbreak among men who have sex with men in Ho Chi Minh City. These models calculated mathematically the level of isolation and social distancing. In fact, defining what activities will reduce 10% or 20% social contacts, what actions will do to isolate infected cases is a matter. This requires consultation of researchers on social science. The mortality rate in our models only covers deaths of COVID-19, but other deaths due to overload of health system. Finally, these models did not consider impacts of the effective medication and vaccine.

## Data Availability

Data used for the modeling is included in the Annex 2 of the manuscript.

## Acknowledgement

We thanks Markus Schwhem and Ha Dao for their comments and contribution in the modeling process, and Duyen Duong for her assistance in preparation of reporting data.

## Authors’ Contributions

TAN developed the modeling method and run the projection, QNC, ALTK, HNN, TNTH contributed to assumptions development and drafted the manuscript.

## Annex 1: Details of the model and its algorithms [3]

Number of susceptible individuals:

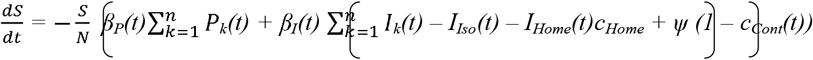

Number of individual s in the latent period:

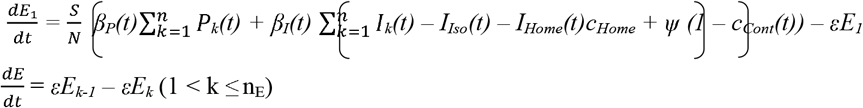

Number of individual s in the prodromal period:

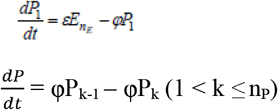

Number of individual s in the symptomatic period:

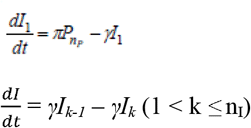

Number of removed individuals:

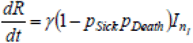

Number of dead individuals:

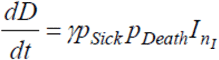

Number of isolated cases at time *t*:

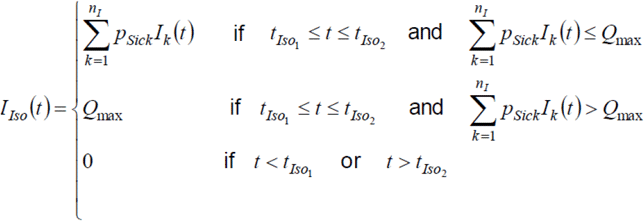

Number of fully isolated cases at time *t*:

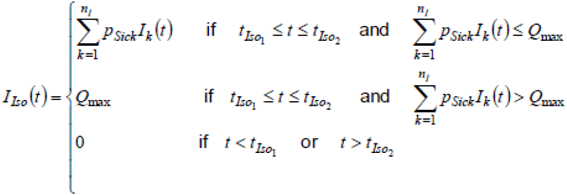

Number of home isolated cases at time *t:*

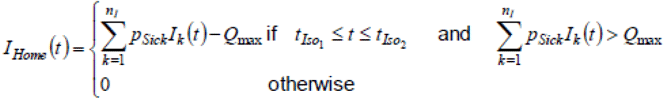

### Initial values

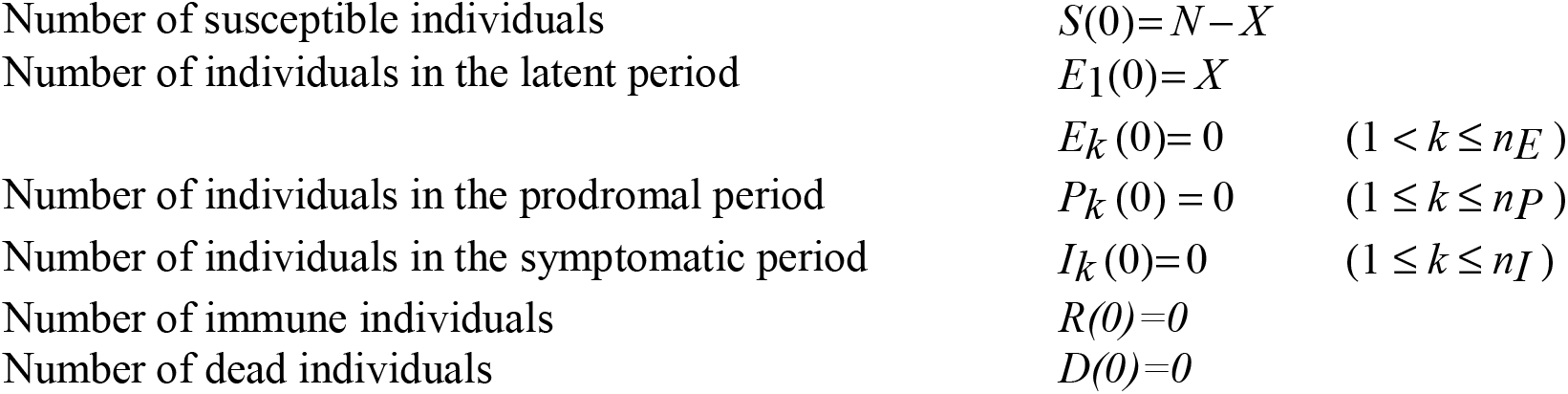

### Parameters

|  |  |
| --- | --- |
| N | Population size |
| X | Number of initial infections |
| $t_{max}$ | Day after introduction of the infection when the transmission potential is highest |
| $Q_{max}$ | Maximum isolation capacity |
| $t_{Iso1}$ | Time at which isolation measures start |
| $t_{Iso2}$ | Time at which isolation measures end |
| $c_{Home}$ | Fraction of contacts which are prevented for cases who are in home isolation |
| $c_{Cont}$ | Fraction of contacts which are prevented |
| $t_{Cont1}$ | Time at which contact reduction starts |
| $t_{Cont2}$ | Time at which contact reduction ends |
| $c(t)$ | Fraction of contacts which are reduced at time $t$ |
| $\psi$ | Force of infection which originates from outside of the population (e.g. via travellers) |
| $R_0$ | Average value of the basic reproduction number |
| $a$ | Amplitude of the seasonal fluctuation of $R_0$ |
| $D_E$ | Average duration of the latent period |
| $n_E$ | Number of stages for the latent period |
| $\varepsilon$ | Stage transition rate in the latent period ($\varepsilon = n_E / D_E$) |
| $D_P$ | Average duration of the prodromal period |
| $n_P$ | Number of stages for the prodromal period |
| $\varphi$ | Stage transition rate in the prodromal period ($\varphi = n_P / D_P$) |
| $i_P$ | Relative infectiousness during prodromal period |
| $D_I$ | Average duration of the symptomatic period |
| $n_I$ | Number of stages for the symptomatic period |
| $\gamma$ | Stage transition rate in the symptomatic period ($\gamma = n_I / D_I$) |
| $\beta_i(t)$ | Effective contact rate of individuals in the symptomatic period at time $t$<br>$\beta_i(t) = R_0 / (i_P D_P + D_I) \cdot (1 + a \cos(t / 365))$ |
| $\beta_p(t)$ | Effective contact rate of individuals in the prodromal period at time $t$ ($\beta_p(t) = \beta_i(t) i_P$) |
| $p_{Sick}$ | Fraction of infected individuals who become sick |
| $p_{Consult}$ | Fraction of sick individuals who seek medical help |
| $p_{Hosp}$ | Fraction of sick individuals who are hospitalized |
| $p_{ICU}$ | Fraction of hospitalized individuals who are admitted to the ICU |
| $p_{Death}$ | Fraction of hospitalized individuals who die from the disease |

### Derived variables

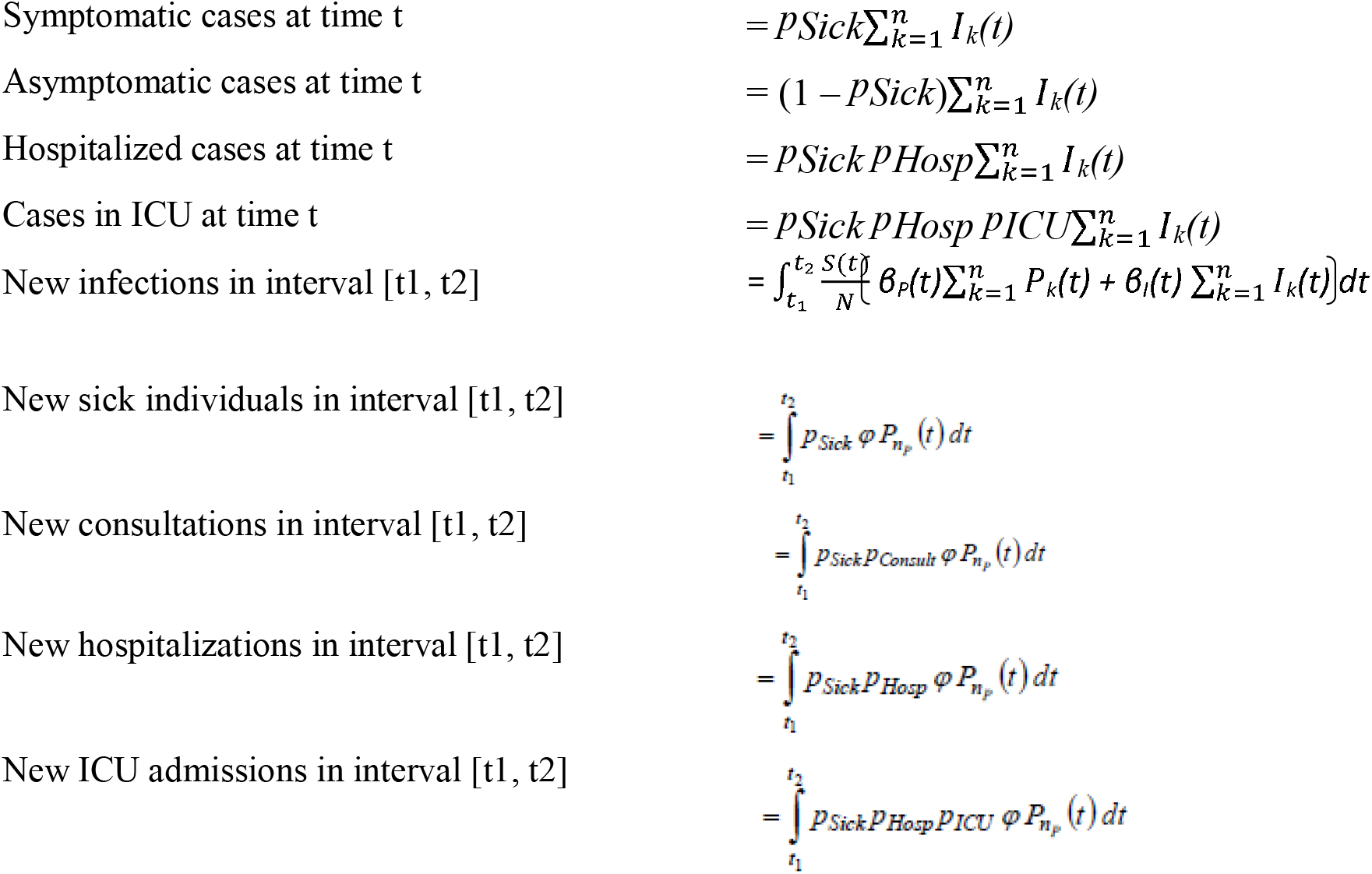
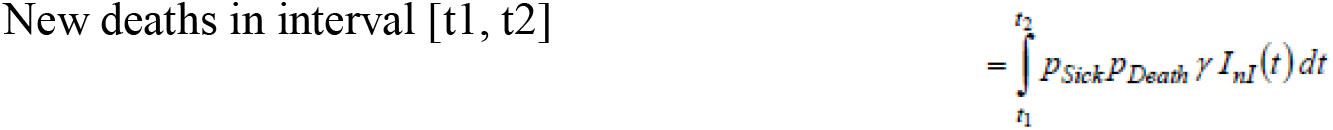

### Detection probability

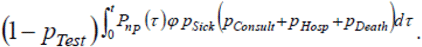

The probability that at least one test has been performed (and has returned a positive result) is then

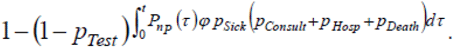

## Annex 2: Input data and assumptions

| Parameter | Value | Details |
| --- | --- | --- |
| **Disease parameters** |  |  |
| **Population** |  |  |
| Population size | 96,208,984 | Source: Viet Nam General Statistic Office [8] |
| Initial infections | 1 |  |
| Infections from outside of the population [per day] | 1 |  |
| **Durations** |  |  |
| Simulation duration [days] | 730 | The simulation is made for a full year, starting from 1 Feb 2020. We assume that the first case found in US by end of Jan and Italy by 3rd week of Feb so it is very likely that the first case entering into Viet Nam is from 1 Feb. This simulation excluded the first cluster of 16 patients who has an epidemiological link to Wuhan. |
| Latency period [days] | 4 | Source: [9] |
| Prodromal period [days] | 1 | Data is not yet available. We use assumption provided by WHO after their first trip to Wuhan and assumption based on data for influenza. Source: [4] [5] |
| Fully infective period [days] | 10 | We assume that 40% of infected cases were notified and 60% were not.<br>Quilty et al. 95%CI: 36-58% [10]<br>Bhatia et al. 95%CI: 63-73% [11]<br>Tuite et al. 95%CI: 61-79% [12]<br>Of those 40% notified, we used the assumption developed based on the WHO-China Joint Mission report which stated that “the median time from onset to clinical recovery for mild cases is approximately 2 weeks and is 3-6 weeks for patients with severe or critical disease”. Assume that 80% of these cases are mild and 20% are severe. The weighted average duration from onset to clinical recovery would be 17.5 days ($80\% \times 14 + 20\% \times 31.5$) [13]<br>Of those 60% not notified, we assumed that the duration from onset to recover would be the minimum estimation in four recent publication: 4.7 days. [14]<br>The weighted average therefore is $60\% \times 4.7 + 40\% \times 17.5 \sim 10$ days |
| Declining period [days] | 5 | Use default value |
| Hospitalization [days] | 15.4 | 13 days [15]<br>7.3 days [14]<br>27 days for 20-29 years old., 32 days for 50-59 years old [16]<br>16.9 days [17]<br>Viet Nam government has policy to keep patients in hospital for quarantine to prevent further transmission.<br>The first 58 patients in Viet Nam: 15.4 days |
| ICU admission [days] | 23.5 | Assume the longest period was found in patients aged 50-59 years old (32 days) [16]<br>Italia: 15 days on average [18]<br>We take the average for the assumption. |
| Number of Erlang stages | 16 | Default setting |
| **Severity** |  |  |
| Infections which will lead to sickness [%] | 82% | Initially 40% of passengers of the Diamond Princess cruise ship, Yokohama did not develop any symptoms; but after end-time follow up, only 17.9% did not develop any symptom. [19] |
| Sick patients seek medical help [%] | 40%<br>60% | Worst case scenario: Default setting, which based on medical consultations for influenza-like illness among people in German from 2017/18.<br>Other scenarios: Number of cases in Viet Nam is too small therefore we cannot estimate the value of this parameter. Patients who have mild symptom would not go to hospital. Patients with severe symptoms accounted for 20% and they are likely to come to hospital. In addition to that, given a consistent communication about COVID-19, we assume that 50% of people with mild symptom would visit health clinic, this proportion among sick population is equal to = 80% mild condition x 50% visiting health clinic = 40%.<br>So in total there is 60% sick people visiting health clinic. |
| Sick patients are hospitalized [%] | 30% | Hospitalization occurred in 30% (13 122 of 43 438) of cases reported from 17 countries (median country-specific estimate, interquartile range (IQR): 24%, 11-41%) [20]<br>In VN this proportion could be lower due to limited criteria for testing. |
| Hospitalized cases need intensive care (ICU) [%] | 10.0% | Severe illness was reported in 15% (1 894 of 12 961) of hospitalized cases from 15 countries (median, IQR: 16%, 10-24%). [20]<br>15.7% patients in severe condition [21]<br>Adjusted for data in Viet Nam: around 10% |
| Sick patients die from the disease [%] | 4.3%<br>3% | Worst case scenario: Meta-analysis (mostly Chinese): 4.3% [22]<br>WHO Situation report 73 dated 2/4: 45,526/896,450 = 5.1%<br>After running the simulation with current policy, the estimated death by Apr 6 is 8, while there is no reported death so far. That is probably because we detected and gave treatment at early stage of infection. There can be death due to COVID-19 but not detected. However, this estimated number is believed to be higher than the actual number.<br>Other scenarios: We adjusted this number to be 3% as the observed death case is still small in Viet Nam. |
| **Contagiousness** |  |  |
| Annual average of the basic reproduction number $R_0$ | 3.4<br>2.5 | Worst case scenario: Use $R_0=3.4$ that was estimated for Wuhan in the beginning of the outbreak<br>Other scenarios: We use $R_0 = 2.5$, using the following evidence Hong Kong and Italy starting Jan 31st till Mar 5: 2.6 [23]<br>WHO: 2 - 2.5<br>A recent review of 12 modelling studies reports the mean $R_0$ at 3.28, with a median of 2.79. [20] |
| Amplitude of the seasonal fluctuation of $R_0$ [%] | 0% | We assume no difference in transmission relate to season. |
| Day when the seasonal $R_0$ reaches its maximum | 0 | We assume no difference in transmission relate to season. |
| Relative contagiousness in the prodromal period [%] | 50% | Use default value |
| Relative contagiousness in the declining period [%] | 50% | Use default value |
| **Detection** |  |  |
| in ILI patients who seek medical help [%] | 0.017<br>0.107 | Worst case scenario: By 20/3 there were 16,000 tests performed (assume one test per one person, although some people got tested several times. However due to lack of the data required, we have to make assumption). Assuming that these tests were used equally used for outpatients, hospitalized and deceased cases, which is 0.017%.<br>Other scenarios: 73164 tests by 2/4 + 30,000 test for planning |
| in hospitalized ILI patients [%] | 0.017<br>0.107 | We assume who were tested are those visited hospital and be hospitalized for isolation. |
| in patients who died from ILI [%] | 0.1 | Use default value |
| **Interventions** |  |  |
| **Case isolation** |  |  |
| Probability that a sick patient is isolated [%] |  | Worse case scenarios: 0%<br>Scenario with 25%, 33%, 50%, 70%, and 80% of sick people isolated.<br>Currently we assume 80% of patients are isolated. Those who are not yet diagnosed or not yet quarantined immediately have a duration of not being isolated. |
| Maximum capacity of isolation wards [per 10,000] | 6<br>11 | Worst case scenario: # of bed per 10,000 was 28 in 2018.<br>Without COVID19, many hospitals are overloaded so we assume only 20% of current beds could be used for COVID-19, or equivalent to 6 beds per 10,000<br>Other scenarios: We added 5 beds/10,000 from quarantine centers |
| Contact reduction for cases in home isolation [%] | 0%<br>50% | Worst case scenario: 0%<br>Other scenarios: We assume that there are undetected patients who have practiced home isolation, and therefore the contact reduction would be assumed as 50%. |
| Begin of case isolation measures [day] | 0<br>33 | Worst case scenario: 0<br>Other scenarios: first case isolated from March 5. |
| Duration of case isolation measures [days] | 0<br>1000 | Worst case scenario: 0<br>Other scenarios: Isolation policy will be applied whenever there is a case. |
| **General contact reduction** |  |  |
| General contact reduction [%] | 0% | Worst case scenario: 5% due to mass media<br>Other scenarios: 10% due to general media |
| Contact reduction begin [day] | 0<br>33 | Worst case scenario: 0<br>Other scenarios: first case isolated from March 5. |
| Contact reduction duration [days] | 0<br>1000 | Worst case scenario: 0<br>Other scenarios: Isolation policy will be applied whenever there is a case. |
| **Triggered General Contact Reduction** |  |  |
| Sick Threshold [%] | 0.021% | We assume that when 200 case was hospitalized, more contact reduction will be practiced by general community. |
| Sick triggered contact reduction [%] |  | Worst case scenario: 5% because some mass communication still has small effect.<br>Other scenarios: 10%, 20%, 25%, 30% (social distancing parameter when more case found) |
| Hospitalization Threshold [per 1.000] | 0.208% | We assume that when 200 case was hospitalized, more contact reduction will be practiced by general community. |
| Hospitalization triggered contact reduction [%] |  | Worst case scenario: 5% because some mass communication still has small effect.<br>Other scenarios: 10%, 20%, 25%, 30% (social distancing parameter when more case found) |
| ICU Threshold [per 100.000] | 2.079% | We assume that when 20 case was in ICU, substantial contact reduction will be practiced by general community. |
| ICU triggered contact reduction [%] |  | Worst case scenario: 5% because some mass communication still has small effect.<br>Other scenarios: 10%, 20%, 25%, 30% (social distancing parameter when more case found) |

## References

[1] W. H. Organization, “Situation report - 79. Coronavirus disease 2019 (COVID-19),” 08 April 2020.

[2] V. M. o. Health, “https://ncov.moh.gov.vn/,” [Online]. Available: https://ncov.moh.gov.vn/.content-type=[Accessed 08 April 2020].

[3] M. S. Martin Eichner, “CovidSIM version 1.1,” University of Tübingen and IMAAC NEXT Association, [Online]. Available: http://covidsim.eu/?fbclid=IwAR1OiZpWVWDZHuIJ5EWA3LLz_CFYBscSocAhy3fdCIKCM9F27LIrI46-08. [Accessed 08 April 2020].

[4] L. T. B. A. K. A. V. M. B. M. S. Nick Wilson, “Modelling the Potential Health Impact of the COVID-19 Pandemic on a Hypothetical European Country,” Preprint, p. https://doi.org/10.1101/2020.03.20.20039776, 2020.

[5] L. T. B. A. K. M. B. Nick Wilson, “Potential Health Impacts from the COVID-19 Pandemic for New Zealand if Eradication Fails:,” University of Otago Wellington, 23/03/2020.

[6] Google, “Vietnam COVID-19 Community Mobility Report (https://www.google.com/covid19/mobility/),” 29 March 2020.

[7] Q. N. C. A. L. T. K. T. N. H. H. N. N. G. J. F. G. B. M. T-A. Nguyen, “Adapting a Tuberculosis contact investigation strategy for COVID-19,” IJTLD, no. https://www.theunion.org/news-centre/news/adapting-a-tb-contact-investigation-strategy-for-covid-19, 2020.

[8] GSO, “Results of the population and housing census 2019 (https://vietnam.unfpa.org/en/news/results-population-and-housing-census-2019),” 2019.

[9] P. S. C. B. S. Y. Z. T. Y. W. e. a. Li R, “Substantial undocumented infection facilitates the rapid dissemination of novel coronavirus (SARS-CoV2).,” Science, 2020.

[10] C. S. C. n. w. g. F. S. E. R. M. Quilty Billy J, “Effectiveness of airport screening at detecting travellers infected with novel coronavirus (2019-nCoV),” Euro Surveill., vol. 25, no. 5 (https://doi.org/10.2807/1560-7917.ES.2020.25.5.2000080), 2020.

[11] https://www.imperial.ac.uk/media/imperial-college/medicine/sph/ide/gida-fellowships/Imperial-College---COVID-19---Relative-Sensitivity-International-Cases.pdf.

[12] V. N. E. R. D. F. Ashleigh R. Tuite, “Estimation of COVID-19 outbreak size in Italy based on international case exportations,” Preprint: https://doi.org/10.1101/2020.03.02.20030049., 2020.

[13] W.-C. J. Mission., “Report of the WHO-China Joint Mission on Coronavirus Disease 2019 (COVID-19).,” 2020 (16-24 February).

[14] A. B. C. A. M. G. G. Z. Alessia Lai, “Early phylogenetic estimate of the effective reproduction number of SARS-CoV-2,” Journal of Medical Viology, no. https://doi.org/10.1002/jmv.25723, 25 February 2020.

[15] Y. D. Z. X. e. a. Penghui Yang, “Epidemiological and clinical features of COVID-19 patients with and without pneumonia in Beijing, China,” https://doi.org/10.1101/2020.02.28.20028068., 2020.

[16] Y. W. S. M. e. a. Qifang Bi, “Epidemiology and Transmission of COVID-19 in Shenzhen China: Analysis of 391 cases and 1,286 of their close contacts,” https://doi.org/10.1101/2020.03.03.20028423., 2020.

[17] J. Y. W. W. e. a. Xiaowei Deng, “Case fatality risk of novel coronavirus diseases 2019 in China,” https://doi.org/10.1101/2020.03.04.20031005., 2020.

[18] K. Z. Davide Manca, “Analysis of COVID-19 data on numbers in intensive care from Italy,” European Society of Anaesthesiology, 2020.

[19] K. K. A. Z. G. C. Kenji Mizumoto, “Estimating the asymptomatic proportion of coronavirus disease 2019 (COVID-19) cases on board the Diamond Princess cruise ship, Yokohama, Japan,” Eurosurveillance, vol. 25, no. 10 (https://doi.org/10.2807/1560-7917.ES.2020.25.10.2000180), 2020.

[20] E. C. f. D. P. a. Control, “Disease background of COVID-19,” 2020.

[21] Z.-y. N. Y. H. e. a. Wei-jie Guan, “Clinical Characteristics of Coronavirus Disease 2019 in China,” New England Journal of Medicine, no. doi:10.1056/NEJMoa2002032, 2020.

[22] S. Q. Z. L. J. R. J. X. Pengfei Sun, “Clinical characteristics of 50466 patients with 2019-nCoV infection,” no. DOI: 10.1101/2020.02.18.20024539.

[23] S. Z. Q. L. Zian Zhuang, “Preliminary estimating the reproduction number of the,” no. https://doi.org/10.1101/2020.03.02.20030312, 2020.

